# Methamphetamine-Contaminated Residences in the United States: Assessment of the Environmental Health Significance of Third-Hand Exposure

**DOI:** 10.1101/2024.07.30.24311229

**Authors:** James E. Dennison

## Abstract

**Objectives:** Methamphetamine (meth) use in the United States has been a significant problem for many years. Beyond impacts to the users, two additional consequences of the meth problem are on-going exposure to non-users in contaminated homes and the significant cost of remediation. This study reports the first U.S. national and state-level estimates of the number of contaminated properties, the number of exposed non-users, and the costs associated with remediation.

**Methods:** The ability to estimate these endpoints relies on having random surveys of the frequency or incidence of residential contamination, but such surveys are difficult to perform and therefore scarce. The results of the only identified random survey were used in this study and appropriate geographical and temporal adjustments were made. The overall rate of contamination of Housing Units (HUs) was determined from estimates of the rates of HU contamination by meth smoking, rates of contamination from meth manufacturing, and rates of HU decontamination. Rates equations were integrated to estimate the number of contaminated HUs, the number of people living in contaminated HUs, and potential remediation costs.

**Results:** The random survey found 3.5% of HUs to be contaminated in 2018. Currently, over 5,000,000 (4%) of U.S. HUs are estimated to be contaminated above average health standards. Based on this, the current estimated cost for remediation of all contaminated HUs would be $15 billion per year and $250 billion for the backlog of all currently contaminated HUs. The estimated number of persons currently exposed to meth above average health standards is 13,000,000 people.

**Conclusions:** While the accuracy of these estimates is limited, they indicate that meth-contaminated housing is a significant environmental health and economic issue in the U.S. that has been previously under-recognized. Additional studies of health effects, fate and transport mechanisms, and remediation methods are needed.

## Introduction

When methamphetamine (meth) is manufactured (“cooked”) or smoked in a residence, the residence readily becomes contaminated with the Schedule II drug (Martyny et al. 2008; VanDyke et al. 2009). As meth has low volatility, it is absorbed onto the building and contents, and occupants experience third-hand exposure dermally, through ingestion, and possibly via inhalation, as the contaminant persists for long periods. Testing and/or remediation may be required if law enforcement apprehends illegal activity involving meth, but many meth labs and houses where meth was smoked (smoke houses) are not identified or tested and no known regular or required general programs of testing exist. Often, testing is conducted when there is suspicion of meth activity (cooking or use), or sometimes just when an owner or other person has a “need-to-know.” However, collections of data from mandated or voluntary tests are expected to be highly biased towards residences at risk of contamination.

Thirteen U.S. states have mandatory cleanup regulations that must be followed if a meth cooking operation (“meth lab”) is found and five other states have requirements that also apply to storage or use (MLCC 2024). USEPA (USEPA 2021) and many other states have voluntary guidelines for cleanup. While some properties are ultimately remediated, others are not due to lack of awareness, resources, concern, requirement/regulation, or other reasons. Meth remediation is usually not subject to government oversight or reporting and is only done if testing is conducted and the presence of meth is found, if at all. Therefore, the number of contaminated residences that became contaminated and have not yet been remediated is unknown.

When testing is conducted, a significant percentage of residences are found to contain traces of meth (below health standards), or to exceed standards. In many cases, these contaminated residences are occupied by non-users. Meth cooking or use results in persistent surface contamination (Bitter 2017) and maybe be airborne as well (Wright et al. 2021). Non-users may occupy during the time meth is used, and receive second-hand exposure, or after use has ceased, and receive third-hand exposure. In many cases, non-users may not be aware of contamination and may in fact first occupy the residence after the use ceased, as in cases when a contaminated residence is purchased, or a new tenant moves into a contaminated apartment.

Most testing is performed at properties where meth activity did or is suspected to have occurred and cannot inform the estimation of actual contamination incidence. Random surveys have not apparently been performed in the past. This is perhaps not surprising as obtaining consent and addressing properties that are found to be contaminated are significant obstacles to performing random surveys. One unique data collection was found, and as previously reported (Dennison and Minick 2024) found that approximately 3.5% of single-family homes and condominiums in the Boulder, Colorado area were contaminated in excess of Colorado’s health standard of 0.5 ug meth/100 cm^2^. In the study performed here we identify the inputs to a cumulative incidence quantity, obtained data supporting these inputs, and thereby calculate estimates of the incidences throughout the U.S.

A previous estimate of meth incidence was reported for Australia (Parker and Howell 2021). The assessment employed meth use data (% of population) and assumed each user would contaminate one HU. The assessment did not ascertain if the user would contaminate the HU to levels above health standards, and no contamination data was involved. The assessment concluded that 1.5% of Australian homes may be contaminated to some degree. The present study seeks to extend this approach by determining the numbers of residences that are expected to exceed health standard, the number of people residing in contaminated units, and the implied remediation costs.

## Methods

A survey of public health officials, consultants who perform meth testing, and meth remediation contractors throughout the U.S. was performed to obtain incidence surveys and remediation cost data. No unbiased or randomized meth surveys were located other than the survey cited here (the Boulder assessment). Property owners and managers were also surveyed for repair cost data. Here, we use the U.S. Census Bureau definition of Housing Unit (HU): “A housing unit is a house, an apartment, a mobile home, a group of rooms, or a single room that is occupied (or if vacant, is intended for occupancy) as separate living quarters. Separate living quarters are those in which the occupants live and eat separately from any other persons in the building and which have direct access from the outside of the building or through a common hall” (U.S.CensusBureau 2024b).

The challenge in finding random meth survey data revolves around the fact that a property is defined as contaminated or not according to comprehensive test protocols that are generally expensive and are generally performed only when evidence of meth activity exists and are therefore biased towards contaminated sites. If limited tests are done in a survey, site selection may still be biased, and sampling error may still occur. The Boulder assessment avoided these challenges to a large extent by using a random screening survey coupled to comprehensive testing conducted for all properties where significant meth was initially detected.

The Boulder assessment was based on data collected by a real estate brokerage (Agents for Home Buyers; (A4HB)) located in Boulder, Colorado. To protect buyers against unwittingly purchasing a property that was contaminated with meth, the broker arranged for buyers to have a meth screening test performed during the inspection period of the real estate contract. 95% of the ∼300 properties that went under contract were screened and most of the non-participants were buying new construction. For houses that were contaminated per the screening, a comprehensive test was typically performed thereafter to confirm whether meth contamination exceeded state health standard. Using the data from the comprehensive follow-up testing, the authors found that 3.47% of the HUs in the survey were contaminated at levels above the Colorado health standard of 0.5 ug meth/100 cm^2^ as of 2018 (Dennison and Minick 2024). This survey result may be considered to be reasonably solid as a minimum estimate because:

1. Relatively little bias occurred in HU selection in the study. Geographical bias is controlled by extrapolating to other states using actual data inputs.
2. All properties were screened with a test protocol that has a very low false negative rate (∼1% or less).
3. The 3.5% estimate ultimately derives from the data from a robust sampling protocol prescribed by Colorado regulations.

Calculations were then performed to translate this into estimates for other years and geographical locations. Other states with regulations have health standard ranging from 0.05 to 1.5 ug/100 cm^2^ (ADEQ 2024; Washington 2024). The calculations described below were run with respect to the entire range of regulatory limits.

### Method for Determining the Total Number of Contaminated Housing Units

The total number of contaminated HUs at a point in time is given by Eq. 1:

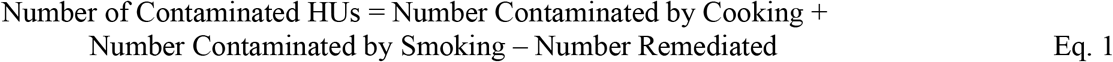

Since we can estimate the number of contaminated HUs using meth use rates, which vary over time and location, and we have access to other rate data, we express this as a rate equation.

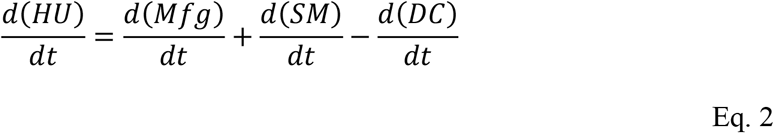

where HU = Contaminated Housing Units

Mfg = Contaminated by manufacturing

SM= Contaminated by smoking

DC = Decontaminated

d/dt = the rate of change in the quantity (HUs/year)

Also, 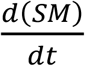can be arrived at as

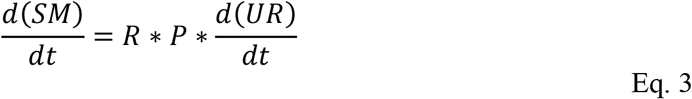

where UR = Use Rate (% of population/year), P = Population, and R is the number of HUs contaminated per user. Published values of R were not found, so were estimated. All other calculation inputs were based on actual data, so the computation contained one degree of freedom, and R could thus be fitted to the result of the published value HU (% contaminated in 2018 in Colorado (3.47%)) using bootstrap methods. With other inputs set to published data, R is thus determined as the number which gives 3.47% of HUs becoming contaminated. Thus, the rate equation can be expressed as Equation 4.

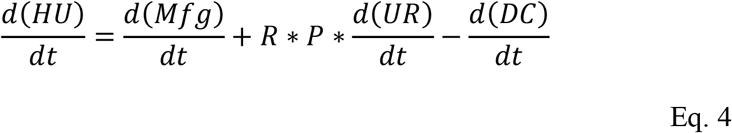

Equation 4 was integrated from 1990 to 2022 to obtain the number of contaminated HUs at any point in time. The rate equation was solved with one-year timesteps in Microsoft Excel. Numerical solution software could be used but Excel was used to facilitate future use of the model by others.

The rate of HUs contaminated by manufacturing (*d(HU)/dt*) in the U.S. and each state was taken from the National Clandestine Laboratory Register Data website (USDEA 2024) which publishes clan lab discoveries from 2004 to present. It is assumed that any amount of manufacturing would result in a contaminated HU based on prior studies (VanDyke et al. 2009), so each discovery was assumed to result in one contaminated HU. While only the majority of DEA database entries were meth-related, there are also likely some unreported. Especially in later years, this variable has a small impact on the overall rate of HU contamination. Individual state and national DEA data was used from 2004 to 2022. Prior to 2004, state and national estimates were estimated as follows. The Department of Justice reported 508 clan labs found nationwide in 1986 (USDOJ N/A) and 6,632 in 2004 and a rapid decline thereafter. We assume that 2004 was the peak year because the Combat Methamphetamine Epidemic Act (CMEA) of 2005 made access to meth precursors substantially more difficult and has been credited with the precipitous decline in meth labs (USDOJ 2006). Each state was assumed to have a pro-rata number of meth labs in 1990 based on population relative to the US population and was extrapolated linearly to 2004.

Use rate data (*d(UR)/dt*) was taken from the annual National Surveys on Drug Use and Health published by the Substance Abuse and Mental Health Services Administration (SAMHSA) (SAMSHA 2013, 2017, 2018, 2019, 2020, 2023). From 2016 to present, state-specific meth use data (% of respondents 12 years or older who used meth in past year) was available except for 2020-2021. Values for 2020-2021 were interpolated. National data was used for the 2002-2022 period for the U.S. rate (SAMSHA 2013). Linear regression was used to predict state values for 2016 (2001 for national), and then was extrapolated from there to a zero rate in 1989.

The rate of meth lab remediation (*d(DC)dt*) was taken from the Colorado Department of Public Health and Environment (CDPHE) database (CDPHE 2024) of post-remediation reports mandatorily submitted by meth consultants for Colorado remedial projects since 2015. Actual remediation data were used for 2015-2022, the average remediations/year for 2015-2022 period was used for 2014 and linear regression of 2015-2022 data was used to estimate to 2005 when Colorado remediation regulations took effect. While other states may have started a program sooner or later than Colorado, voluntary remediations were occurring even prior to 2005, so a non-zero value is justified. For 1990 to 2004, a linear extrapolation to zero was then used. As a small fraction of remediations are performed in property other than housing, this rate is a slight overestimate. For the U.S. and other states, the Colorado rate was extrapolated by the ratios of housing units in Colorado to other states in 2022.

The total number of housing units in the U.S. and states were from 2022 (U.S.CensusBureau 2024b), prorated based on population in 1990 and linearly extrapolated between 1990 and 2022. Actual census data for population was used for all years (Macrotrends 2024).This data was compiled into an Excel file with 52 worksheets (U.S., 50 states, D.C.) and is available online in Supplemental Materials. Integration of the rate equation was conducted with one-year timesteps starting in 1990 and the total contaminated HU quantity was aggregated from the prior year’s total plus the total of new contaminated HUs.

As discussed below, the A4HB survey found 3.47% of HUs tested between 2013-2022 were contaminated with meth. As the % of HUs contaminated increases over time, it was assumed that the rate of 3.47 was reached in the middle of this survey period, *i*.*e*., in 2018. The value of R (#HUs contaminated per user) was determined by adjusting the value of R until the % of Colorado HUs that were contaminated was ∼3.47% in 2018. This R value was then used for the U.S. and all states.

### Cost data

The costs associated with meth-contaminated properties fall into three categories:

1. “Remediation costs” are the costs of returning a building to a defined non-contaminated state. Occasionally, buildings are simply demolished, but a high percentage are remediated. Removal of materials such as carpet, cabinets, and drywall is common, or cleaning of any or all surfaces may be done. Remediation cost means the money paid to the remediation contractor to return the house to compliance. This cost includes associated re-testing costs.
2. Repair costs include replacement of all items that were removed during remediation. These commonly include carpet, appliances, fixtures such as lights, re-painting, re-insulating, replacement of wall or ceilings, HVAC systems, and other items.
3. Other costs include such items as loss of use, management time, loss of equity due to stigmatization, legal fees, and other costs.

Based on surveys of contractors and consultants, a reasonable average cost for meth remediation across all types of properties is $20,000 per housing unit. The cost for studio apartments, one-bedroom apartments, or other smaller spaces would normally be lower. Some buildings may be contaminated only in selected parts, as little as perhaps one room, and would cost less than the average. However, a whole house will typically cost more than $20,000, depending on the size, degree of contamination, mobilization costs, amount of occupant contents, and the chance of concomitant asbestos, lead, or other abatement work necessitated to remove contaminated building materials. In our experience, remedial costs have ranged from $5,000 to $45,000 per housing unit.

Repair costs and other costs likewise range broadly. Loss of use is usually for a period of 3-6 months. Based on the survey, the average cost for replacement and other costs has been $29,000 for single family homes and $15,000 for condominiums or apartments. An average of $24,000 per unit will be used in this assessment. Thus, the total cost for remediation, repair and other costs is estimated at $44,000 per unit on average.

The Colorado estimate was based directly on the 3.47% HU contamination rate in 2018. Other states and the U.S. as a whole were adjusted for state-specific use data, meth lab DEA registry data, populations and housing data, using the value for R determined from Colorado.

## Results

In the A4HB survey (Dennison and Minick 2024), 288 housing units were screened for methamphetamine over the ten-year period and 35 housing units were also tested with a comprehensive test. Ultimately, 10 of the original 288 housing units (3.47%) were found to be contaminated above Colorado’s health standard. According to the screening data, 12.5% of housing units in the area contain traces of meth less than the health standard of 0.5 ug/100 cm^2^, and 84% contain no detectable meth.

Table 1 provides the state-by-state and national breakdown of calculation results, including use rates, #HU contaminated per year, Total HUs currently contaminated, % of all HU contaminated, total remediation costs, and # of persons living in contaminated units. The data shown are for 2022; earlier years can be viewed in Supplemental Materials in the state tabs. In Colorado, the % HU contaminated rose from ∼3.5% in 2018 to 4.1% as of 2022 due to the low remediation rate compared to the rates of contamination. The U.S. and other states % HU contaminated is mostly related to the relative Use Rates, shown in Table 1.

**Table 1:** Current Estimates of Contaminated Units, Remedial Costs and Total Exposed Persons.

| Location | Regulation Status | Cleanup Level,<br>ug/100 cm2 | Use Rate in<br>Last year, % | HU<br>Contaminated<br>per Year | Total HU<br>Contaminated | % of all HU<br>contaminated | Cleanup Cost<br>Million\$\$ | Persons living in<br>Contaminated<br>HUs |
| --- | --- | --- | --- | --- | --- | --- | --- | --- |
| United States | Guidelines | None known | 0.95 | 350581 | 5,671,801 | 3.94% | \$ 249,559 | 13,146,207 |
| Alabama | None known | None known | 1.40 | 7962 | 99,355 | 4.25% | \$ 4,372 | 215473 |
| Alaska | Cook only | 0.10 | 1.01 | 821 | 15,539 | 4.72% | \$ 684 | 34603 |
| Arizona | Rescinded | None known | 0.84 | 6807 | 169,889 | 5.33% | \$ 7,475 | 392689 |
| Arkansas | Cook only | 0.05 | 1.74 | 5983 | 55,768 | 4.00% | \$ 2,454 | 121724 |
| California | Cook or storage | 1.5 | 1.35 | 59393 | 669,305 | 4.58% | \$ 29,449 | 1786372 |
| Colorado | Cook or use | 0.50 | 0.81 | 5187 | 107,089 | 4.13% | \$ 4,712 | 241299 |
| Connecticut | None known | None known | 0.41 | 1544 | 22,998 | 1.49% | \$ 1,012 | 53878 |
| Delaware | None known | None known | 0.70 | 774 | 10,595 | 2.27% | \$ 466 | 23189 |
| District of Columbia | None known | None known | 0.39 | 263 | 4,040 | 1.12% | \$ 178 | 7521 |
| Florida | None known | None known | 0.53 | 12503 | 170,615 | 1.66% | \$ 7,507 | 370016 |
| Georgia | None known | None known | 0.81 | 9736 | 105,781 | 2.33% | \$ 4,654 | 254322 |
| Hawaii | Cook only | 0.10 | 0.91 | 1453 | 29,616 | 5.21% | \$ 1,303 | 75042 |
| Idaho | Cook or storage | 0.10 | 0.86 | 1370 | 25,540 | 4.50% | \$ 1,124 | 64713 |
| Illinois | None known | None known | 0.93 | 12938 | 62,509 | 1.15% | \$ 2,750 | 144244 |
| Indiana | Cook only | 0.50 | 1.24 | 9476 | 102,068 | 3.43% | \$ 4,491 | 234224 |
| Iowa | None known | None known | 1.27 | 4544 | 51,293 | 3.57% | \$ 2,257 | 114087 |
| Kansas | None known | None known | 0.88 | 2850 | 67,328 | 5.21% | \$ 2,962 | 152963 |
| Kentucky | Cook only | 0.10 | 1.67 | 8496 | 93,289 | 4.61% | \$ 4,105 | 207972 |
| Louisiana | None known | None known | 1.62 | 8368 | 38,453 | 1.82% | \$ 1,692 | 83485 |
| Maine | None known | None known | 1.05 | 1605 | 10,534 | 1.40% | \$ 463 | 19467 |
| Maryland | None known | None known | 0.41 | 2640 | 31,235 | 1.22% | \$ 1,374 | 75232 |
| Massachusetts | None known | None known | 0.51 | 3786 | 43,455 | 1.43% | \$ 1,912 | 99934 |
| Michigan | Guidelines | None known | 0.62 | 6695 | 79,557 | 1.73% | \$ 3,501 | 173088 |
| Minnesota | Cook or use | 0.1/1.5 | 1.04 | 6597 | 93,614 | 3.67% | \$ 4,119 | 209949 |
| Mississippi | None known | None known | 1.54 | 5090 | 41,582 | 3.10% | \$ 1,830 | 91006 |
| Missouri | None known | None known | 1.38 | 9559 | 115,680 | 4.09% | \$ 5,090 | 252813 |
| Montana | Cook only | 1.5 | 1.52 | 1917 | 19,679 | 3.72% | \$ 866 | 41760 |
| Nebraska | Cook only | 0.10 | 1.07 | 2341 | 21,184 | 2.45% | \$ 932 | 48259 |
| Nevada | Unknown | Unknown | 2.23 | 8054 | 56,885 | 4.28% | \$ 2,503 | 136030 |
| New Hampshire | Unknown | 0.10 | 0.64 | 966 | 16,414 | 2.53% | \$ 722 | 35407 |
| New Jersey | None known | None known | 0.74 | 7512 | 39,097 | 1.03% | \$ 1,720 | 95652 |
| New Mexico | Cook only | 1.0 | 1.56 | 3710 | 42,405 | 4.43% | \$ 1,866 | 93671 |
| New York | None known | None known | 0.50 | 10427 | 73,104 | 0.85% | \$ 3,217 | 167500 |
| North Carolina | Cook only | 0.10 | 0.69 | 7998 | 124,538 | 2.55% | \$ 5,480 | 272254 |
| North Dakota | None known | None known | 1.18 | 1022 | 16,380 | 4.34% | \$ 721 | 33773 |
| Ohio | None known | None known | 1.11 | 14519 | 104,306 | 1.97% | \$ 4,589 | 231725 |
| Oklahoma | None known | 0.10 | 1.41 | 6369 | 78,214 | 4.40% | \$ 3,441 | 176940 |
| Oregon | Cook only | 0.05 | 1.18 | 5583 | 95,333 | 5.13% | \$ 4,195 | 217358 |
| Pennsylvania | None known | None known | 0.62 | 8668 | 131,746 | 2.27% | \$ 5,797 | 293879 |
| Rhode Island | None known | None known | 0.70 | 832 | 9,283 | 1.91% | \$ 408 | 20891 |
| South Carolina | None known | None known | 1.07 | 6271 | 54,688 | 2.24% | \$ 2,406 | 118082 |
| South Dakota | Guidelines | 0.10 | 1.05 | 1060 | 13,112 | 3.21% | \$ 577 | 29218 |
| Tennessee | Cook only | 0.10 | 1.41 | 11156 | 95,360 | 3.03% | \$ 4,196 | 213759 |
| Texas | None known | None known | 0.82 | 27165 | 288,656 | 2.38% | \$ 12,701 | 714222 |
| Utah | Cook only | 1.0 | 1.16 | 4403 | 45,773 | 3.73% | \$ 2,014 | 125958 |
| Vermont | None known | None known | 0.52 | 352 | 8,102 | 2.39% | \$ 357 | 15464 |
| Virginia | None known | None known | 0.42 | 3809 | 65,701 | 1.78% | \$ 2,891 | 154728 |
| Washington | Cook or Storage | 1.5 | 1.25 | 10895 | 106,629 | 3.22% | \$ 4,692 | 250438 |
| West Virginia | Cook only | 1.0 | 2.20 | 4423 | 36,468 | 4.23% | \$ 1,605 | 75085 |
| Wisconsin | None known | None known | 0.71 | 4534 | 77,187 | 2.79% | \$ 3,396 | 164107 |
| Wyoming | Cook only | 0.75 | 1.43 | 932 | 7,081 | 2.56% | \$ 312 | 14863 |

The U.S. and selected states use rates are shown in Figure 1. The national use rate (0.95%) for 2022 is similar to Colorado (0.81%) and thus the % contaminated in the U.S. is also similar (3.94%).

**Figure 1:**
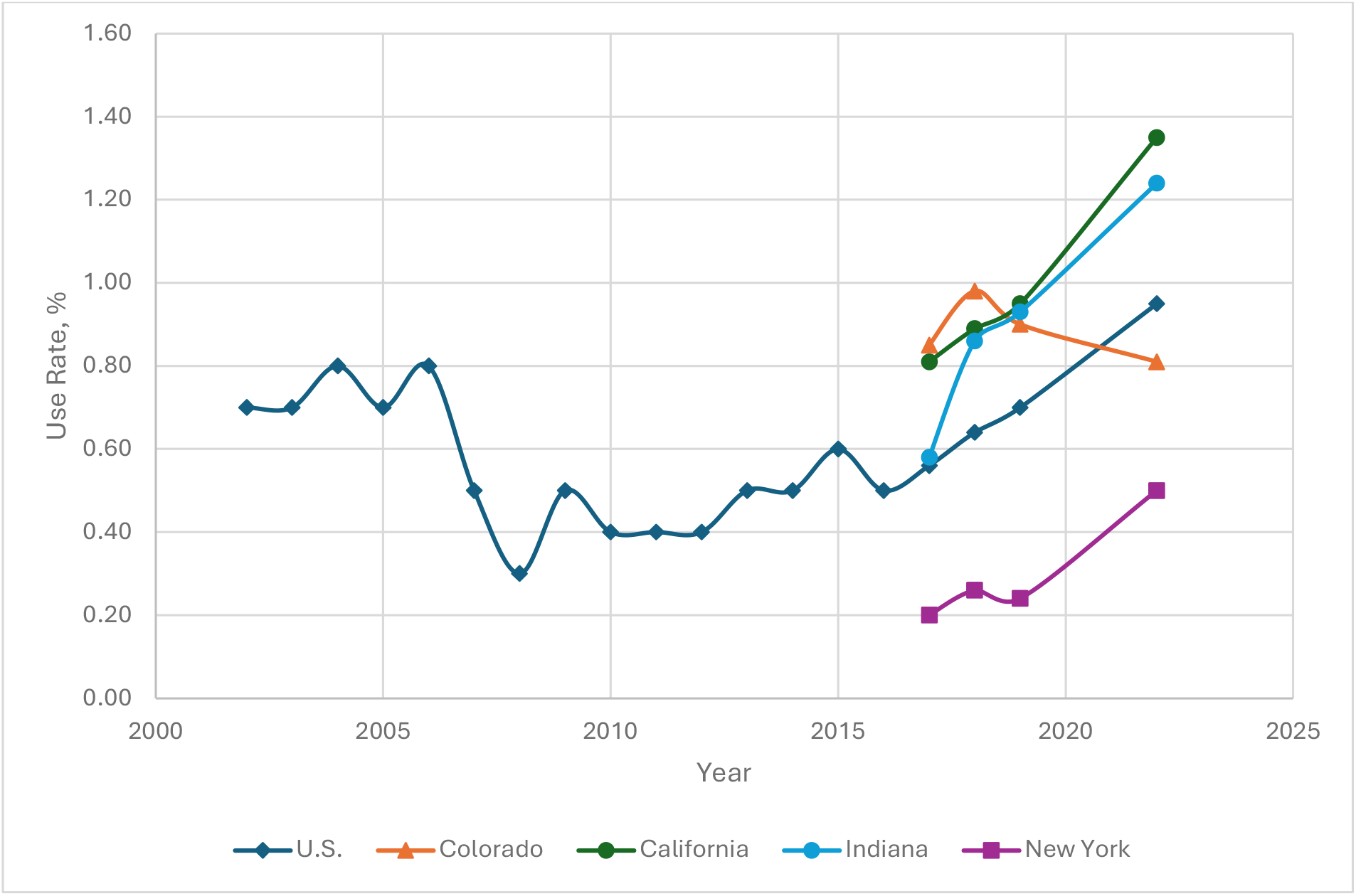
Meth Use Rate in U.S. and Selected States (Age 12+ in past year (%))

The data provided in Table 1 is based on the Colorado health criterion (0.5 ug/100 cm^2^.) Based on this criterion, the U.S. is estimated to have over 5 million contaminated HUs, and over 13,000,000 people exposed above the regulatory limit with a cost of $250 billion to remediate. The number of U.S. HU contaminations per year and the aggregate number is shown in Figure 2. The annual cost to the U.S. economy for newly contaminated building remediation would be approximately $15 billion. State-specific estimates are also provided. According to the A4HB data, 16% of all HUs contained detectable meth at concentrations above or below the health criterion. In the U.S., 22 million people would be exposed to meth at any detectable concentration.

**Figure 2:**
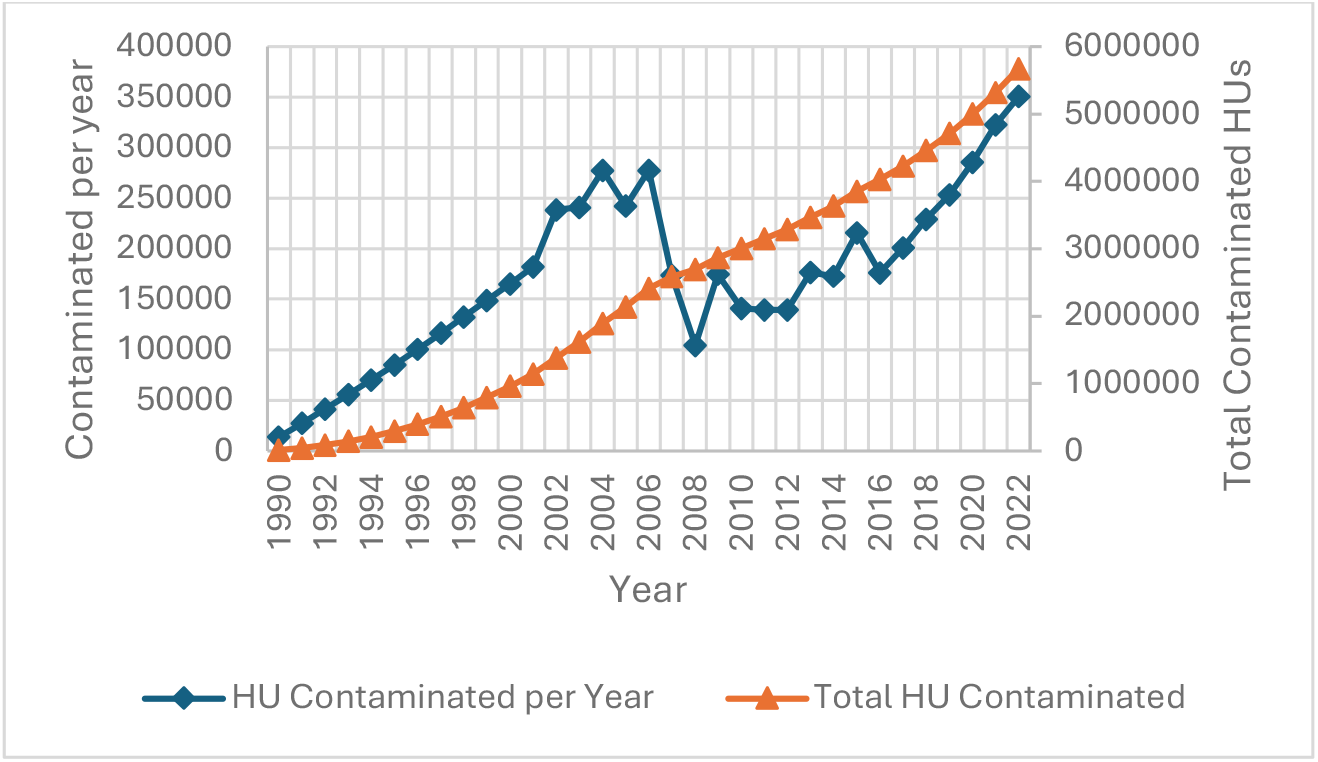
HU Contaminations in U.S.

In Colorado, the rate of new contaminations is currently approximately 5,000 HUs per year. Data from mandatory reporting to CDPHE of all compliance testing and post-decontamination reports shows that approximately 200 HUs are remediated per year, about 4% of the estimated number created. The fraction remediated in other states is typically lower than in Colorado based on interviews of professionals in other states. While the 2018 estimates should be as accurate as the A4HB survey data is, the estimated number of newly created contaminated HUs for subsequent years may be overestimated. This would result from the tendency for users to contaminate the same HU from year to year, and for the contamination of the same units by different persons with no remediation in between. Extrapolating the calculated results much past 2022 would not be reliable.

Meth use data was taken from the on-going SAMHSA National Survey on Drug Use and Health. The agency did not report the data from 2020-2021 due to changes in methodology and states that 2022 is not comparable to earlier years. Indeed, the rates in 2022 appear to be higher than before. Nevertheless, rates in many states appear to be rising from 2017 to 2019 as well, and the U.S. rate has been increasing since 2010 (Fig. 1). While the rate of increase in contaminated HUs is uncertain, it seems likely that the incidence will increase if use rates increase or remain at present levels and decontamination rates remain low.

Unlike use, the incidence of meth lab (manufacturing operations) is decreasing, very significantly after 2005, with many states currently reporting no meth lab discoveries in a typical year (Figure 3).

**Figure 3:**
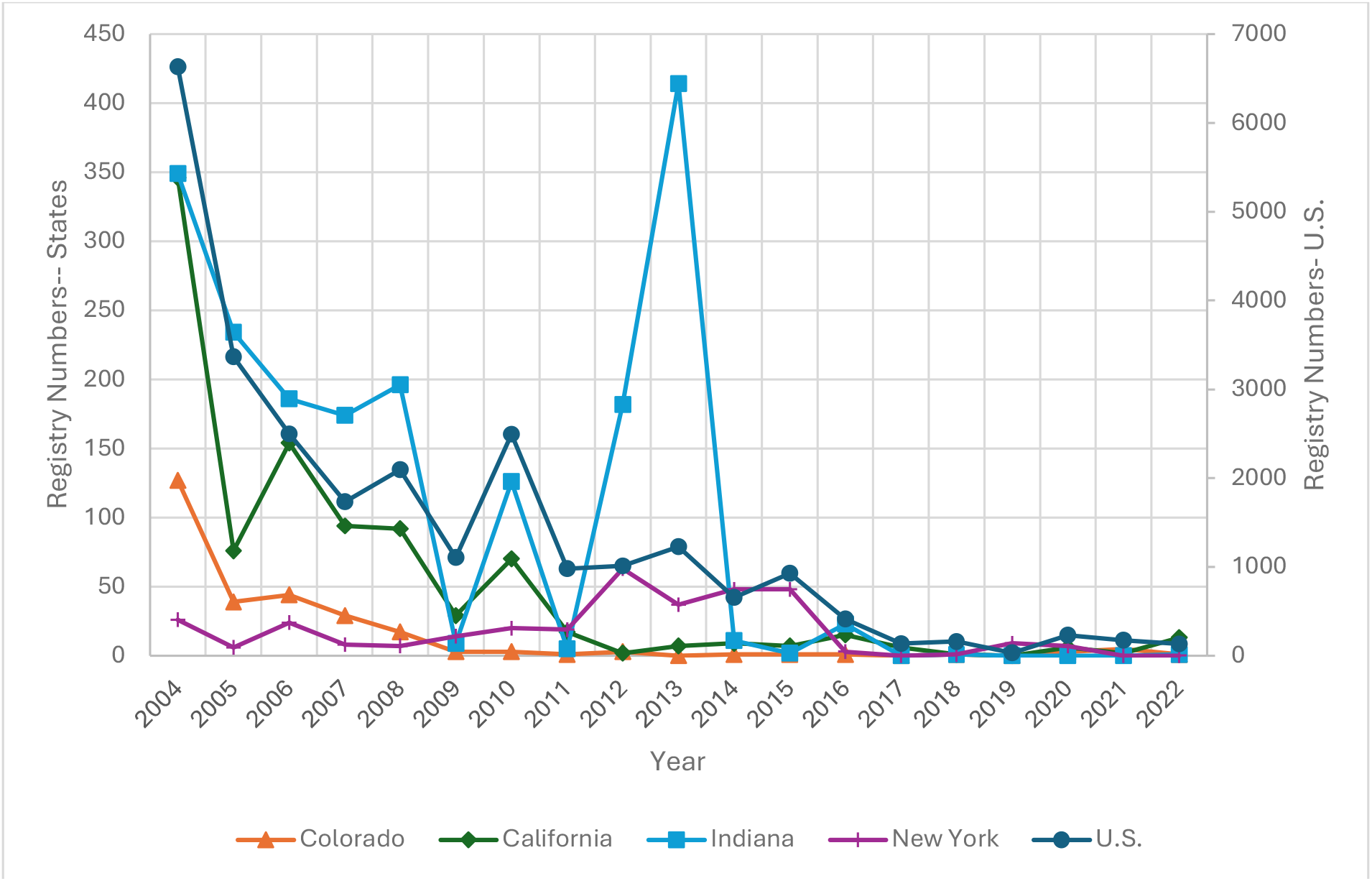
DEA Clan Lab Registry numbers

Various states have promulgated health criteria ranging from 0.05 to 1.5 ug/100 cm^2^. As previously reported (Dennison and Minick 2024), higher criteria result in fewer contaminated HUs, lower aggregate costs, and fewer people exposed above standards. The decrease was more prominent at first (at lower standard levels) and then the rate of reduction declines. At the highest published health criterion of 1.5 ug/100 cm^2^, over 3 million HUs are estimated to be contaminated, occupied by almost 8 million people. These are lower estimates than when the Colorado criterion is used, but obviously still substantial.

## Discussion

This analysis of the only identified random survey of meth incidence data suggests that there is a substantial number of housing units in the United States that are contaminated with meth. Thus, there is a very substantial aggregate cost for cleanup of these units, with more contamination on-going, and a very substantial number of individuals exposed to meth in a third-hand manner at concentrations above state health standards.

Many people living in contaminated housing are unaware of the meth in their residence. Contamination is sometimes present when the person(s) moves into the home, or they may not be aware of meth use when it occurs. Other times, they might be aware of its use, but don’t realize that the home can become contaminated from use. Relatively few homes are ever tested. The result of this knowledge gap is that numerous housing units remain contaminated and cause third-hand exposure. If health effects can indeed occur at concentrations above health criteria, this is a significant environmental health concern.

### Uncertainty Analysis

This assessment used data that was not a direct measure of the endpoints. Use rates underlie *d*(SM)*dt*, DEA Registry data were a surrogate for *d*(Mfg)/*dt*, and Colorado remediation reports represented *d*(DC)*dt*. In general, extrapolations in parameter values were made with the “lowering result” assumption. For example, lacking systematic data for remediation rates in other states, the Colorado rate was used for the entire U.S. after adjustment for housing stock even though remediation rates are likely lower in states with no or cooking-only regulations. The intent was to err on the side of estimating low but not grossly so.

As the value for R was obtained using the 3.47% outcome of the A4HB survey, this parameter is the main source of uncertainty, and all state and national contamination rates fall in a range close to this value. The uncertainty is the Boulder rate was discussed elsewhere (Dennison and Minick 2024), but includes the following:

1. The population of HUs in the survey was in and around Boulder and Boulder County, Colorado but was extrapolated to the entire state. Boulder and Boulder County are above average in various measures of socioeconomics, including income, housing costs and education, which tends to suppress drug use (Johnston 2006; Tejiram et al. 2022; U.S.CensusBureau 2024a). Other data support the idea that Boulder may have a lower incidence than Colorado at large. While meth use data for the Boulder area was not identified, SAMHSA reported that cocaine use was 24% lower in the Boulder area than in Colorado at large (SAMHSA 2019).
2. The 3.47% rate did not include some properties that tested positive in the screening and did not undergo a comprehensive test. This may have reduced the rate by ∼25%.
3. The A4HB survey did not include apartments, and rarely included rental property which are at higher incidences of contamination than owner-occupied HUs.

Thus, the A4HB survey is more likely to be biased low than high and the estimates for other states and the U.S. would be concomitantly lower than actual.

There is also uncertainty in the estimate of remediation and other costs associated with remediation. A random survey for this data was not conducted as part of this study. Based on specific examples we reviewed, the cost estimates are accurate to at least +/− 50% and for the purposes of obtaining an order of magnitude estimate of costs, the approach appears adequate. Regardless of this uncertainty, the results show that remediating all contaminated housing units would bear a very significant cost. The severe burden this places on property owners is predictable. The effect on low-income rental housing (agency or non-profit corporation) is that the total cost for dealing with meth contamination has been said to decimate maintenance and improvement budgets, and essentially forces the policy that testing cannot practically be done unless an absolute mandate exists, as most units tested are found to be contaminated (Gonzales 2024). This approach, despite being a result of economic imperatives, results in unrecognized third-hand exposures. As this study does not address other types of buildings or vehicles, or other drugs often also present (e.g. fentanyl), the cost of the generalized problem is yet higher.

### Why Are So Many Housing Units Contaminated?

It may seem surprising that so many housing units are contaminated with meth, but an understanding of the process and rate of contamination provides insight. The contaminant level in a building is the net of introduction and elimination processes.

As shown, meth use has increased while meth manufacturing has dramatically declined in the U.S. Thus, current contamination of buildings is almost entirely due to use (primarily smoking). The present analysis indicates that, since 1990, only approximately 1% of HUs were contaminated by cooking, and the current fraction is only 0.04% due to the success of the CMEA. Data on the amount of contamination that occurs during a single use of meth in real-life is not currently available but might be estimated. Consider smoking a typical amount (0.2 g) of meth in a bedroom: if 1% of the meth in the original product is released from the pipe and user, in a typical bedroom of dimensions 4m x 5m x 3m, a one-compartment no-ventilation model surface concentration would become 0.2 ug/100 cm^2^. This calculated estimate is supported by Martyny et al. (Martyny et al. 2008) who measured meth concentrations after a simulated “smoke” in the range of 0.2-0.3 ug/100 cm^2^. It has been reported that sampling after a single suspected use of meth in buildings showed contaminant levels close to the detection limit of 0.1 ug/100 cm^2^. Absent elimination, the contaminant level is additive after each use. Thus, after several uses, a building would reach levels defined as contaminated.

The crystalline form of meth is methamphetamine hydrochloride. After smoking, methamphetamine vapor, the hydrochloride form, and combustion by-products are present. Sample analysis will yield a combined result for all forms of meth. The melting point for meth is 170 °C and the boiling point is 212 °C (PubMed 2024) and the hydrochloride form would be higher yet, placing meth into the semi-volatile organic compound class. Thus, at ambient room temperatures, meth and its hydrochloride would coalesce onto building surfaces in a solid state and would be adsorbed by dust and other particles. With an estimated vapor pressure of 0.005 mm Hg at 25 °C, (NCBI 2024) sublimation or evaporation is a slow process. Within buildings, contamination appears to persist for a long time with highly elevated concentrations found even years after the activity ceased.

Due to the low vapor pressure, most meth in buildings is expected to be absorbed by surfaces and particles (technically, some may be adsorbed, and the rest absorbed, but the exact mechanisms are not well understood at this time). Indeed, the risk assessment performed by Cal EPA (CalEPA 2009a) assumed that exposure to humans in contaminated buildings would be through dermal absorption and ingestion, and inhalation would not be significant. However, Wright et. al. (Wright et al. 2021) conducted a study concluding that inhalation may be significant and deserves further study. These exposures to meth in contaminated buildings have been called third-hand smoke (Yeh et al. 2022) or here, third-hand exposure.

Very little is known about the fate and transport of meth in buildings and is an area in need of further investigation. Yet, with low vapor pressure, losses due to evaporation should be slow. *In-situ* degradation is possible, but no data is currently available. Other removal methods such as ordinary cleaning are usually applied to small portions of the total building, so accumulation over years does indeed makes sense. Thus, with use rates of ∼1% of the population, single use contaminant levels estimated at 0.2 ug/100 cm^2^, and additive accumulation, a 3.5% incidence of contaminated housing units appears plausible.

### Why is Contamination from Manufacturing a Small Part of the Problem?

The CMEA required that pseudoephedrine (PSE), ephedrine, and phenylpropanolamine be placed “behind the counter” and that transaction logs account for sales of these substances from retail outlets. Ephedrine and PSE are precursor chemicals used in the production of methamphetamine. In the CMEA, law enforcement officials were authorized to have access to these logs, and retail outlets were allowed to sell only 3.6 grams of PSE a day per individual purchaser. There appears to be a correlation between the passage of the CMEA and a decrease in meth cooking in the United States, perhaps aided by restrictions of other chemicals needed for meth production and enhanced law enforcement. Over the range of years for which data were available, meth use has significantly increased, with imported methamphetamine filling the demand gap. Thus, the rate of building contamination is not anticipated to decrease in the near future. Even in the early days of meth regulation, the environmental health issue was never really limited to HUs contaminated by cooking. While there were additional chemical contaminants that might have existed, they typically were never sampled, and were assumed to be remediated if the meth was remediated. Thus, the regulations that require remediation only when meth was cooked may be addressing less than 1% of the problem defined by health standards.

Increasingly, other drugs such as fentanyl have also become a concern. Use of other controlled substances such as cocaine and heroin is common (CDC 2022) but started long ago, and always presented the possibility that a HU could be contaminated with more than one drug. A newer trend is for some drugs (e.g., meth) to be cut with another (e.g. fentanyl), so more than one would tend to be in a HU.

### Effect of Alternate Health Criteria

The concept of “contaminated” is definition dependent. As previously mentioned, states with regulatory limits have promulgated criteria between 0.05 and 1.5 ug/100 cm^2^, a thirty-fold range. At this time, USEPA has not promulgated a standard or provided a numeric guideline (USEPA 2021). In the A4HB survey, if the criterion adopted by Oregon and Arkansas (0.05 ug/100 cm^2^) was used, 8% of housing units would be deemed contaminated. If the California criterion of 1.5 ug/100 cm^2^ criterion is used, only 2% of housing units would have been deemed contaminated. The incidence of contaminated units is also dependent on the test protocol. If four single surface samples are collected in a room rather than one four-part composite sample and the highest result is used in the data interpretation, more units would be deemed contaminated. Which regulatory limit is appropriate is a matter of debate, but the large difference in standards underscores the paucity of data needed to perform risk assessment. As with any standard, the uncertainty results in either too many or too few properties being remediated, with public health or cost impacts.

### Uncertainty in Risk

Two environmental risk assessments have been performed to date for meth-contaminated buildings. The first published risk assessment was performed by the state of Colorado (CDPHE 2005). In the literature review, no human toxicity data for the target population were found. The risk assessment considered various short-term rodent reproductive and developmental studies.

Available neurological studies in rodents were limited to 20 days duration. The Colorado risk assessment concluded that the proposal for a limit of 0.5 ug/100 cm^2^ would not lead to excess exposure relative to the studies considered, with a typical safety margin added. California EPA performed a risk assessment with a more detailed exposure model to derive their standard (CalEPA 2009b, 2009a). All animal data were considered, but the most pertinent study found for the risk assessment was a human study in which pregnant women were administered low doses of meth to assist with weight control (Chapman 1961). The use of a therapeutic endpoint study in environmental health risk assessment is highly unusual. Along with detailed exposure modeling, CalEPA derived a criterion of 1.5 ug/100 cm^2^, thirty times higher than the lowest published limit, which was not based on risk assessment. The absence of long-term bioassays, human behavioral or cognitive effect studies, neurodevelopmental studies, or other studies after chronic thirdhand exposure remains a concern for a neuroactive agent primarily found in residences.

### Uncertainty in Remediation Techniques

Improvements in remediation techniques can often be best developed by the private sector. However, there are sometime barriers to progress. In some jurisdictions, the use of oxidants is discouraged or banned due to lack of knowledge on oxidative chemistry ^(CDPHE 2014)^. According to USEPA guidance: “The interaction of bleach and meth is not fully understood, and their by-products are currently unknown. <u>Until further research is conducted</u> to identify these by-products and their health effects, bleach should not be used as a cleaning agent….” (USEPA 2021) (emphasis added). At remediation costs of ∼$20K per unit and ∼5 million affected properties, progress on this topic would be timely. The effectiveness of encapsulation, which is also sometimes banned (CDPHE 2014), may also be worthy of study. Serrano et al. initiated studies that showed short term effectiveness of encapsulation (Serrano et al. 2012), but these studies have yet to be conducted over relevant time periods. As meth use is an on-going problem, advances in making remediation less costly are imperative.

At this time, it may be possible to say that there is more we don’t know about third-hand meth exposure than we know. Obviously, there is a profound need to conduct long term or other studies in both animals and humans to further explore the risks of third-hand exposure to meth given the large number of exposed humans. Such studies will hopefully build confidence in the risk assessment and allow better consensus on health criteria. We also have significant knowledge gaps concerning the fate and transport of meth in buildings, which leads into challenges on remediation. For many other environmental health issues in the indoor environment, such as asbestos in building materials, the remediation cost is essentially a one-time cost, but for meth, the problem and costs are recurring. The lack of knowledge regarding effective remediation techniques is a challenge, or perhaps an opportunity, to enhance public health, if remediation costs can be brought into an affordable range rather than leaving contaminated housing units in their present state.

## Data Availability

All data produced in the present study are available upon reasonable request to the authors

## Acknowledgements

Helpful comments were provided by Tim Lockhart, CIH and Colleen Brisnehan.

The author declares he has no competing interests but is engaged in meth consulting activities.

